# Platelet transcriptomic signatures in pediatric brain tumors distinguish cancer from cancer-free control

**DOI:** 10.64898/2025.12.17.25341356

**Authors:** Markus Talka, Amanda Holmström, Jonathan Hiekka, Matti Kankainen, Satu Långström, Mia Westerholm-Ormio, Anu M. Suominen, Ruth Nousiainen, Pirjo Isohanni, Pia Valle, Elina Välkesalmi, Oskar Saijonmaa, Atte Karppinen, Päivi Koroknay-Pal, Anna Piippo-Karjalainen, Jarno Satopää, Nuutti Vartiainen, Jukka Kanerva, Olli Tynninen, Soili Kytölä, Anna-Kaisa Anttonen, Virve Pentikäinen, Katja Eloranta

## Abstract

**Background:** Malignant pediatric brain tumors remain the leading cause of cancer-related mortality in children. Current diagnostics rely on imaging and invasive biopsy, which may not capture tumor heterogeneity. Liquid biopsy-based biomarkers such as tumor-educated platelets have shown diagnostic value in adult cancers, but their utility in pediatric brain tumors has not been investigated.

**Methods:** We analyzed platelet transcriptomes of 73 blood samples from 23 pediatric brain tumor patients, classified as high-grade tumor patients or low-grade tumor patients, and 25 cancer-free controls. Platelets were isolated, CD45⁺ depleted, and RNA was extracted for RNA sequencing. CD45⁺ depletion efficiency was assessed using xCell-based leukocyte enrichment scores. Differential gene expression was assessed with DESeq2 and Gene Ontology over-representation analysis. Gene-level discrimination between groups was evaluated by receiver operating characteristic analysis, and a logistic regression model with patient-grouped 5-fold cross-validation was trained to classify high-grade tumor patients vs. controls.

**Results:** Platelets from brain tumor patients showed transcriptional remodeling compared to controls, especially pronounced in high-grade tumor patients. We identified 398 and 649 differentially expressed genes in the brain tumor group vs. controls and high-grade tumor patients vs. control comparisons, respectively, and 85 genes in high-grade tumor patients vs. low-grade tumor patients. No genes met significance in low-grade tumor patients vs. controls. High-grade tumor patients showed consistent upregulation of cancer-associated mitochondrial genes. Similarly, gene enrichment analyses highlighted pathways related to mitotic regulation, chromosome segregation, and mitochondrial metabolism. Multiple genes demonstrated strong diagnostic performance, and logistic regression classifier based on selected platelet transcripts achieved an area under the curve of 0.89, sensitivity of 79%, and a specificity of 80% in identifying high-grade tumor patients.

**Conclusions:** This study provides the first evidence that platelets exhibit distinct transcriptomic signatures in pediatric brain tumor patients. Platelet RNA profiles robustly differentiate high-grade tumor patients from low-grade tumor patients and cancer-free controls, reflecting tumor presence and biological aggressiveness. These findings support the feasibility of tumor-educated platelets as a minimally invasive biomarker for pediatric malignant brain tumor detection, and longitudinal monitoring. Larger multicenter studies are warranted to validate applicability.

## Background

Around 400,000 children and adolescents under 19 are diagnosed with cancer each year (1). Malignancies affecting the central nervous system (CNS) are the most common solid pediatric malignancies, representing about one-third of all cases. They differ markedly from adult brain tumors in histology and biology (2). The main pediatric brain tumor types include gliomas, medulloblastomas, ependymomas, and germ cell tumors (3), with the highest incidence between ages 0–6 years (4). Despite advances in surgery and therapy, brain tumors account for the highest mortality among childhood cancers (5). Prognosis varies widely: some low-grade tumors have >90% 5-year survival, whereas non-infant high-grade gliomas remain associated with poor outcomes (around 45–50% survival) (6). This highlights the need for accurate diagnosis to avoid overtreatment, and to ensure that aggressive tumors receive timely, intensive management.

The diagnostic gold standard for pediatric brain tumors is surgical biopsy followed by histological and molecular profiling (7), but this approach has notable limitations. Biopsies may miss intra-tumoral heterogeneity (8), and in some cases safe access is not possible. Repeated biopsies are not feasible for follow-up, which instead relies on imaging and cerebrospinal fluid (CSF) cytology, methods that lack molecular resolution. Early biological changes often occur before tumors are radiologically detectable (9,10), and MRI reveals only macroscopic lesions without capturing molecular alterations relevant for progression, relapse, or targeted therapy. Distinguishing true progression from pseudo-progression (e.g., radiation necrosis) is also difficult and may lead to overtreatment (11). These challenges highlight the need for complementary diagnostic tools that enable precise, longitudinal monitoring and earlier detection of tumor development, treatment resistance, or relapse.

Liquid biopsies have recently emerged as an increasingly prominent area of investigation, leveraging tumor-derived biomolecules and tumor-host interactions measured from bodily fluids, such as blood or CSF (9,12). This diagnostic approach enables safer and more frequent follow-up, and it may better capture tumor heterogeneity and minimal residual disease cost-effectively (13,14). Within this landscape, circulating platelets have gained growing interest as a biologically active and readily accessible compartment with diagnostic potential (15). Platelets are small anucleate fragments derived from megakaryocytes. Although they lack nuclei, they retain and translate RNA. Their RNA repertoire reflects megakaryocyte biology but is continuously reshaped by platelet interactions with other cells and systemic physiological cues. Previous studies indicate that malignant processes can influence platelets to acquire tumor-related transcriptomic profiles, giving rise to so-called tumor-educated platelets (TEPs) (16–18). Direct interactions between tumor cells and platelets occur by receptor binding, resulting in tumor-induced platelet activation. Additionally, activated platelets can absorb circulating biomolecules, such as proteins, nucleic acids, and extracellular vesicles from other blood or immune-related cells within the tumor microenvironment (17,19). The uptake of these tumor-derived and host-derived biomolecules reprograms platelet RNA content and function, forming the basis for their designation as TEPs. These altered platelets have been implicated in several pro-tumorigenic processes, including facilitating angiogenesis and supporting metastatic dissemination (20–22).

Taken together, the dynamic remodeling of platelet RNA content and the 10-day platelet lifespan enable platelet transcriptomes to serve as sensitive, near–real-time indicators of disease progression, tumor recurrence, or treatment. Several studies have demonstrated that TEP profiling enables accurate monitoring of disease status across multiple malignancies, including adult glioblastoma, non-small cell lung cancer, colorectal cancer, pancreatic cancer, breast cancer, and hepatobiliary cancers (18). TEP-based assays have shown strong performance in distinguishing progression from pseudo-progression, monitoring actual tumor burden, and recognizing disease relapse (11). These findings highlight the substantial diagnostic and clinical utility of TEPs as a minimally invasive, sensitive, and precise tool for longitudinal cancer surveillance.

To the best of our knowledge, TEPs have not been studied in the context of pediatric brain tumors previously. This study aimed to characterize tumor-induced alterations in platelet gene expression in a pediatric brain tumor patient cohort. Secondly, our objective was to assess the potential of platelet transcriptome signatures in pediatric brain cancer diagnostics.

## Patients and methods

### Sample collection and platelet isolation

Samples were collected in collaboration with Helsinki Biobank. A total of 73 blood samples (4 – 10 mL each) from 23 pediatric brain tumor patients (BTG) and 25 pediatric patients with no known cancer history (control group (CTG)), were collected into EDTA-tubes at the New Childreńs hospital between August 2023 and November 2024 (Helsinki, Finland). Among the 23 brain tumor patients, all had a baseline sample, either at primary diagnosis or at relapse, collected before the treatment initiation. Of these, 9 also underwent serial sampling (2–9 samples per patient). Tumors were classified by experienced neuropathologists using the 2021 World Health Organization CNS tumor classification system (23). Based on the classification, we divided brain tumor patients into two groups: high-grade tumor patients (HGT) and low-grade tumor patients (LGT).

All samples were processed within 2 hours of collection. The whole blood samples were centrifuged at 120 g x 20 min at room temperature (RT) to separate the platelet-rich plasma (PRP). The PRP was then centrifuged at 360 g x 20 min (RT) to pellet the platelets (18). The platelet pellet was then resuspended in solution containing phosphate-buffered saline, pH 7.2, 0.5% bovine serum albumin, and 2 mM EDTA. Subsequently, Miltenyi MACS separator was used with human CD45⁺ microbeads (cat. 130-045-801) and LD columns (cat. 130-042-901; all from Miltenyi Biotec, Germany) for additional leukocyte depletion. The CD45⁺ depleted platelets were pelleted by centrifugation and resuspended in pre-chilled 1-Thioglycerol-Homogenization solution (cat. AS1390, Promega, Madison, Wisconsin) and stored in –80 **°** C.

### Clinical data

Information about diagnoses, tumor grading, received treatments, and proliferation index (Ki-67) were acquired from electronic health records and were processed pseudonymized for study purposes. Additionally, age at sample collection and biological gender were recorded.

### RNA extraction, library preparation, and sequencing

The RNA was extracted from platelet pellets using Promega Maxwell SimplyRNA cells kit (cat. AS1390, Promega, Madison, Wisconsin). The total RNA quality and quantity were assessed using Agilent 2100 Bioanalyzer RNA 6000 Pico kit (cat. 5067-1513, Agilent Technologies, Santa Clara, California). A total of 1000 picograms of high-quality platelet RNA was used for RNA sequencing library preparation using SMARTer Stranded Total RNA-Seq Kit v3 Pico (cat. 634490, Takara Bio USA, Inc., San Jose, California). In cases where less than 1000 picograms was available, the maximum obtainable amount of RNA was used. The quality and quantity of the final libraries were assessed with Agilent 2100 Bioanalyzer DNA high sensitivity kit (cat. 5067-4626, Agilent Technologies, Santa Clara, California) and KAPA Library Quantification kit (cat. 07960204001, Roche, Basel, Switzerland). Finally, libraries were pooled and sequenced using Illumina Novaseq 6000 systems (Illumina, San Diego, California) for 2 x 100 paired-end-sequencing. On average, we generated 42.0 million reads (21.0 million paired-end read-pairs) per sample with an average mapping rate of 96.5%. The base quality along the reads was high, with an average of 96.5 % of bases exceeding the Q30-score.

### Data processing and differential gene expression analysis

FASTQ files were generated from raw base-call .bcl files using Illumina bcl2fastq2 (v.4.3.13). SortMeRNA (v.2.1b) was used to remove rRNA reads from FASTQ files. Trimmomatic (v.0.39) was then used to remove adapter sequences, discard short reads, and trim reads based on base quality scores. Only read pairs in which both mates survived were retained for further processing. The processed high-quality reads were mapped to the human reference genome (GRCh38) using the STAR aligner (v.2.7.10a), with gene annotation from Ensembl release 111. Gene expression quantification was performed using the Rsubread package (v. 2.4.3) within the R statistical environment. The featureCounts tool was utilized to count reads mapping to exons with the following parameters: GTF.featureType = exon, GTF.attrType = gene_id, allowMultiOverlap = TRUE, strandSpecific = 2, isPairedEnd=T, and minMQ=255. Differential gene expression (DE) analysis was performed using the DESeq2 R package (v. 1.48.1) based on raw count data. Genes with a false discovery rate (FDR) < 0.05 and log₂ fold change > 1 or < -1 were considered significantly differentially expressed.

### XCell cell type enrichment analysis

For the xCell cell type enrichment analysis (24), raw gene-level counts were normalized to reads per kilobase per million (RPKM) using gene lengths from the matching Ensembl gene annotation and library sizes. The resulting RPKM matrix was uploaded to the xCell web application (https://comphealth.ucsf.edu/app/xcell) using xCell gene signatures N = 64, which returns enrichment scores for immune and stromal cell types. For downstream immune-focused analyses, the xCell matrix was restricted to a prespecified CD45⁺ cell-type panel with 33 cell types from xCell data: **B cells** (B-cells, Class-switched memory B-cells, Memory B-cells, naive B-cells, pro B-cells); **CD4 T cells** (CD4⁺ T-cells, CD4⁺ Tcm, CD4⁺ Tem, CD4⁺ memory T-cells, CD4⁺ naive T-cells, Th1 cells, Th2 cells, Tregs); **CD8 T cells** (CD8⁺ T-cells, CD8⁺ Tcm, CD8⁺ Tem, CD8⁺ naive T-cells); **γδ T cells** (Tgd cells); **NK/NKT** (NK cells, NKT); **dendritic cells** (DC, aDC, cDC, iDC, pDC); **monocytes/macrophages** (Monocytes, Macrophages, Macrophages M1, Macrophages M2); **granulocytes** (Neutrophils, Eosinophils, Basophils); and **mast cells** (Mast cells). A composite CD45⁺ score was computed per sample as the sum of the xCell scores. Group differences between CD45⁺ depleted and non-depleted samples were tested with a two-sided Wilcoxon rank-sum test. For visualization, a heatmap of the CD45⁺ panel was generated by z-scoring each cell type across samples, followed by hierarchical clustering using Euclidean distances.

### Gene ontology analyses

Over-representation analysis for Gene Ontology (GO) Biological Process terms using the most significant genes (top 20) from filtered DESeq data (FDR < 0.05 and log₂FC > 1 or < -1), was performed using the clusterProfiler package with visualization supported by enrichplot (25,26). Gene annotation was based on org.Hs.eg.db (27). The analysis included the top genes from both comparisons (HGT vs. LGT and HGT vs. CTG). The same genes were visualized as violin plots for both group comparisons. Additionally, the same analyses were performed for all differentially expressed genes from the BTG vs. CTG comparison, with the same filter criteria.

### Statistical analyses

All further statistical analyses were performed using R version 4.3.0. All figures and visualizations were created in R, using packages including ggplot2, readr, dplyr, ggrepel, stringr, tibble, tidyr, rlang, pheatmap, glmnet, pROC, and EnhancedVolcano. Statistical significance is denoted in figures as follows: *p* < 0.05 (*), *p* < 0.01 (**), *p* < 0.001 (***), and *p* > 0.05 (NS; not significant).

Welch’s two-sample t-test was used to assess the group differences in RNA concentrations per 1 mL of plasma between brain tumor patients and cancer-free controls. Pearson’s and Spearman’s correlations were computed to evaluate the association between total platelet count and RNA concentration per sample. A linear regression model (RNA concentration ∼ total platelet count) was fitted to estimate slope and coefficient of determination (R²).

Effect sizes were defined as logFC for overlapping genes to capture magnitude and direction across comparisons, and absolute values were plotted to assess concordance. Gene-level discrimination was evaluated by ROC analysis using log-transformed counts, with AUC calculated by trapezoidal integration. Sample-wise expression of ROC genes was visualized in heatmaps after row-wise z-scoring.

Binary classification assessed whether selected TEP marker genes discriminate HGT from CTG. RNA-seq data were log₁₀(1+CPM) normalized, filtered to CTG/HGT samples, and matched to marker genes (AUC ≥ 0.80). A patient-grouped 5-fold cross-validation was used with z-scoring, zero-variance filtering, and class-weighting. An elastic-net logistic regression model (α = 0.5) generated test-set P(HGT) at lambda.min, and combined predictions yielded ROC AUC, sensitivity, specificity, and accuracy. Gene importance was derived from coefficients of a full-dataset elastic-net model at the optimal penalty.

## Results

### Characteristics of the study population

Plasma samples were collected from 23 pediatric patients with brain tumors (biological gender: 14 male, 9 female; median age at diagnosis 9 years, range 0–17) and 25 cancer-free controls (biological gender: 14 male, 12 female; median age at sample collection 11 years, range 1–17). Among the patients, 15 were diagnosed with high-grade and 8 with low-grade brain tumors based on the assessment of tissue biopsy by histology and molecular profiling or imaging, and CSF cytology results in non-feasible cases where biopsy was not obtained. Among the most common diagnoses were high-grade gliomas, medulloblastomas, low-grade gliomas, and ependymomas. A total of 73 plasma samples were obtained and out of these, 23 samples were collected as follow-up samples during therapy. Demographic and clinical characteristics of the study cohort are provided in **Table 1**. **Table 2** presents a summary of sample collection timepoints and amounts of collected samples at each timepoint. The overall study workflow is summarized in **Figure 1**.

**Figure 1.**
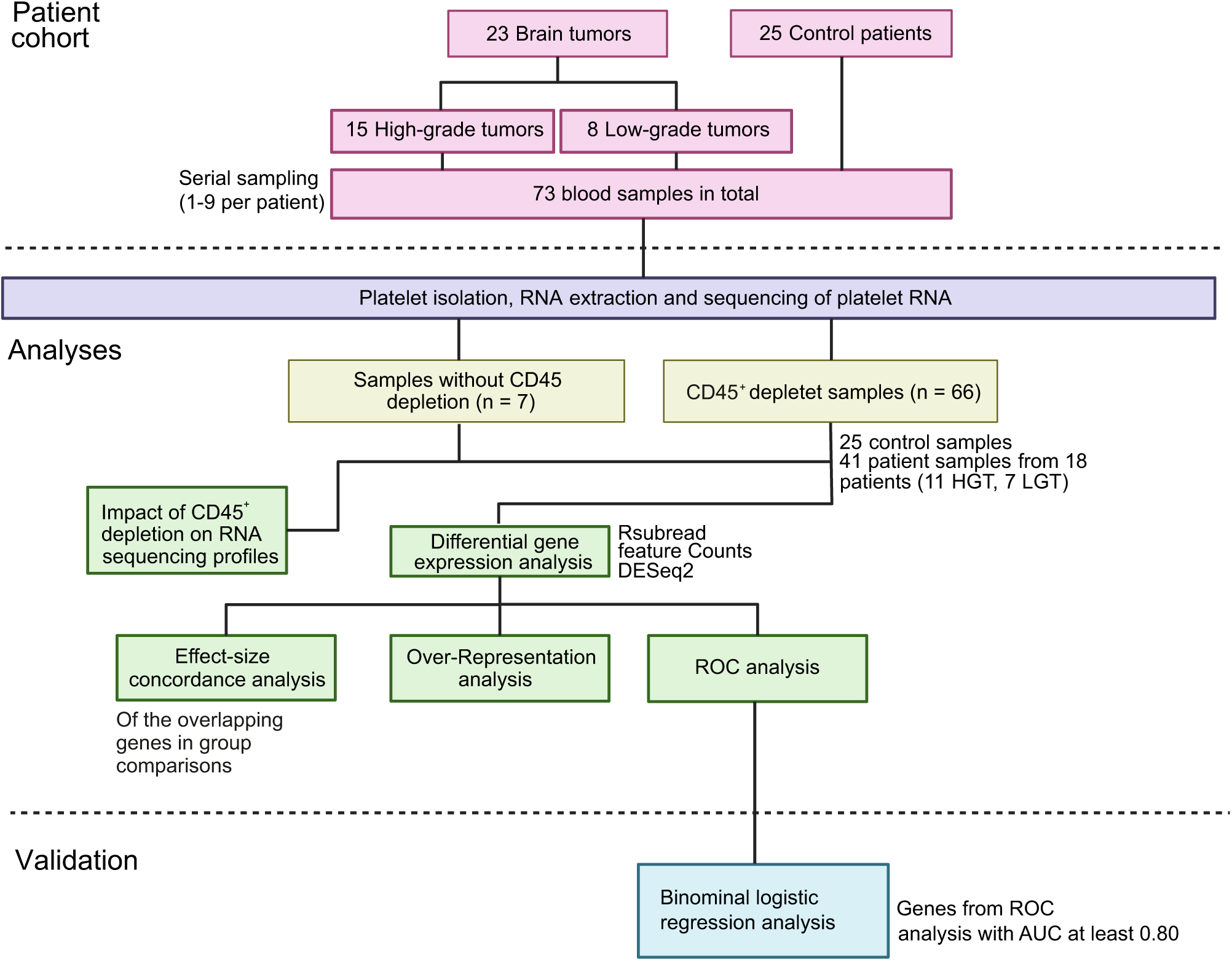
Study workflow. Overview of the experimental and analytical pipeline.

**Table 1.** Clinical characteristics of study patients. Summary of pediatric brain tumor patients including demographics, diagnosis (WHO classification), tumor grade, Ki-67 index and treatments.

| Study ID | Biological gender | Age group at first sample collection, years | Diagnosis | Tumor grade | Ki-67 index (%) | Treatments |
| --- | --- | --- | --- | --- | --- | --- |
| BT102 | Male | 5–10 | Diffuse midline glioma H3K27M altered, Grade 4 | High-grade | 5 | CT, RT, TT |
| BT104 | Male | 11–15 | Medulloblastoma, Group 4, Grade 4 | High-grade | 25 | TR, CT, RT |
| BT105 | Male | ≥ 16 | Medulloblastoma, Group 4, Grade 4 | High-grade | 20 | PR, CT, RT, TT |
| BT106 | Male | 5–10 | Pilocytic astrocytoma, Grade 1 | Low-grade | 6 | TR |
| BT107 | Male | 5–10 | Medulloblastoma, Group 4, Grade 4 | High-grade | 50 | TR, CT, RT |
| BT108 | Female | 5–10 | Diffuse intrinsic pontine glioma* | High-grade | NA | CT, RT |
| BT109 | Male | 1–4 | Pilocytic astrocytoma, Grade 1 | Low-grade | 7 | CT, TT |
| BT110 | Female | ≥ 16 | Pilocytic astrocytoma, Grade 1 | Low-grade | 2 | TR |
| BT111 | Male | 1–4 | Pilocytic astrocytoma, Grade 1 | Low-grade | 3 | CT, TT |
| BT112 | Male | 5–10 | Diffuse pediatric-type high-grade glioma, H3-wildtype and IDH-wildtype, Grade 4 | High-grade | 10 | CT, RT, TT |
| BT113 | Female | 11–15 | Bithalamic low-grade glioma** | Low-grade | 1 | Follow-up |
| BT114 | Female | 11–15 | Low-grade glioma** | Low-grade | 3 | PR, CT |
| BT115 | Female | 5–10 | Pilocytic astrocytoma,<br>Grade 1 | Low-grade | 2 | TR |
| BT116 | Female | < 1 | Atypical<br>teratoid/rhabdoid tumor,<br>Grade 4 | High-grade | 90 | CT, SCT |
| BT117 | Male | 5–10 | Diffuse midline glioma<br>H3K27 altered, Grade 4 | High-grade | 20 | CT, RT, TT |
| BT118 | Male | 11–15 | Diffuse pediatric-type<br>high-grade glioma,<br>Grade 4 | High-grade | 85 | CT, RT |
| BT119 | Male | 5–10 | Germinoma, Grade 4 | High-grade | 90 | CT, RT |
| BT120 | Female | 1–4 | Posterior fossa group A<br>(PFA) ependymoma,<br>Grade 3 | High-grade | 8 | PR, CT, RT |
| BT121 | Female | 11–15 | Supratentorial<br>ependymoma ZFTA<br>fusion-positive, Grade 3 | High-grade | 20 | TR, RT |
| BT122 | Male | 11–15 | Diffuse pediatric-type<br>high-grade glioma,<br>Grade 4 | High-grade | 55 | CT, RT, IT |
| BT124 | Male | 5–10 | Pilocytic astrocytoma,<br>Grade 1 | Low-grade | 2 | TR |
| BT125 | Female | 5–10 | Germinoma, Grade 4 | High-grade | 95 | CT, RT |
| BT126 | Male | 11–15 | Supratentorial<br>ependymoma ZFTA<br>fusion-positive, Grade 3 | High-grade | 50 | TR, CT, RT |

Treatment abbreviations: CT = chemotherapy, RT = radiotherapy, TT = targeted therapy, IT = immunotherapy, SCT = autologous stem cell transplantation, TR = total resection, PR = partial resection. \* = no biopsy was obtained. \*\* = the histopathology did not match any WHO diagnosis; alternative diagnosis was suggested.

**Table 2.**
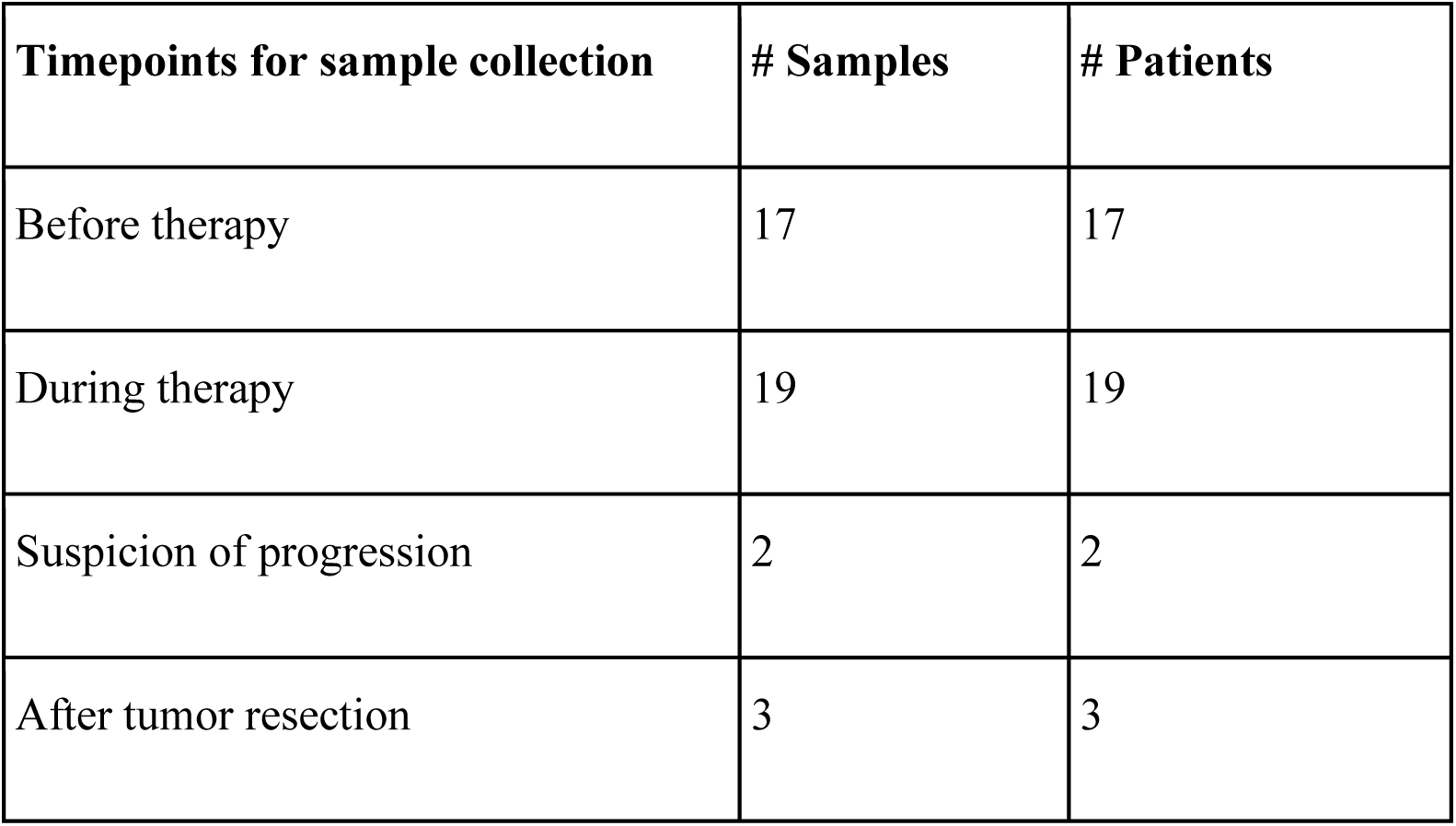
Summary of longitudinal platelet samples from brain tumor patients. The table presents the total number of samples and individual patients for each collection stage. The stages correspond to: **Before therapy:** samples collected before therapy initiation, usually on the day of biopsy; **During therapy:** samples collected while patients were undergoing treatment; **Suspicion of progression:** samples collected at suspicion of disease progression, either during or after therapy; **Resected:** samples collected after tumor resection, either partial or complete.

| Timepoints for sample collection | # Samples | # Patients |
| --- | --- | --- |
| Before therapy | 17 | 17 |
| During therapy | 19 | 19 |
| Suspicion of progression | 2 | 2 |
| After tumor resection | 3 | 3 |

### RNA extraction, quality assessment, and correlation with platelet count

RNA was successfully isolated from all plasma samples and evaluated for yield and integrity. CD45⁺ depleted samples (n = 66) yielded a mean plasma volume of 2.3 ± 0.8 mL (range 0.9–4.0), versus 1.8 ± 0.5 mL (range 1.0–2.5) in non-depleted samples (n = 7). Median RNA concentration was 592.5 pg/µL (Q1–Q3: 134.2–1714.2; range: 7–8840) in the depleted group and 2913 pg/µL (Q1–Q3: 2610–6225; range: 1300–9620) in the non-depleted group. Corresponding means were 1426.3 ± 1927.0 pg/µL and 4500.4 ± 3472.6 pg/µL, respectively. RNA integrity (RIN) was high in both groups: RIN 8.9 ± 0.7 (range 6.4–10.0) in the depleted group and 8.1 ± 0.8 (range 6.8–9.4) in the non-depleted group.

Although RNA yield varied between samples, the extracted RNA quantities were sufficient for all subsequent analyses. To account for differences in plasma input volume, RNA concentrations were normalized to 1 mL of collected plasma prior to comparison. For the comparison, we included only CD45⁺ depleted samples with available plasma input volume (n = 61). No significant difference was observed between BTG and CTG in RNA concentration (p = 0.71) (**Figure 2A).**

**Figure 2.**
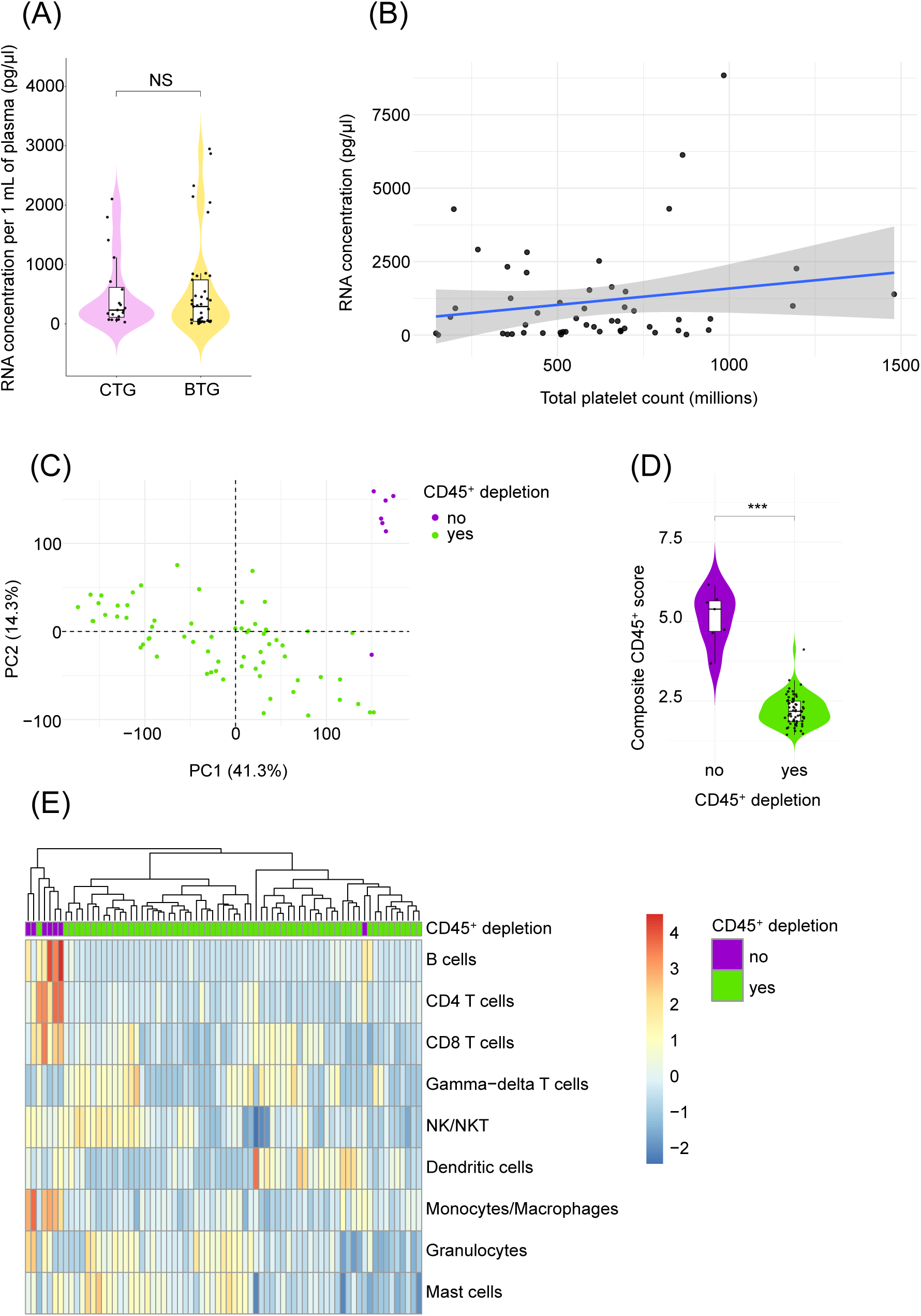
Platelet RNA extractions and correlations between BTG and CTG, and the impact of CD45⁺ leukocyte depletion on sample purity. **(A)** Violin plot of normalized RNA concentrations in 1 mL of plasma. **(B)** Scatterplot of correlations between extracted RNA concentrations and total platelet counts. BTG = brain tumor group; CTG = cancer-free control group. NS = statistically not significant. **(C)** PCA plot from CPM normalized RNA seq data shows clustering of CD45⁺ depleted and non-depleted samples based on transcriptomic profiles. **(D)** Violin plot of the composite CD45⁺ xCell scores shows enrichment of CD45⁺ cells in samples without CD45⁺ depletion. **(E)** Clustering heatmap visualization of different CD45⁺ cell types across samples. Most of the samples without CD45⁺ depletion cluster together. Rows depict cell types and columns depict samples. *** = p-value < 0.001.

Correlation analyses between RNA concentration and platelet count were restricted to CD45⁺ depleted samples (n = 51 after excluding samples without platelet counts). Platelet count showed no meaningful association with plasma RNA concentration. A simple linear model explained 3.4% of the variance (R² = 0.034), and correlations were weak and non-significant (Pearson r = 0.183, p = 0.198; Spearman ρ = 0.194, p = 0.172). As shown in **Figure 2B**, datapoints were broadly dispersed with no monotonic trend across platelet counts.

### CD45⁺ depletion reduces leukocyte contamination in samples

To evaluate the CD45⁺ depletion efficiency, a subset of samples did not undergo CD45⁺ depletion prior to RNA extraction. Principal component analysis (PCA) of the RNA sequencing data revealed that non-depleted samples clustered separately, indicating that depletion affected overall transcriptomic profiles (**Figure 2C)**. To further investigate the CD45⁺ depletion efficiency, a composite CD45⁺ depletion score was calculated based on xCell scores by summing the enrichment scores across a predefined set of leukocyte signatures and compared between the CD45⁺ depleted (n = 66) and non-depleted (n = 7) samples. The CD45⁺ depleted samples showed a markedly lower composite score (median 2.171) than non-depleted samples (median 5.386), corresponding to an approximate 60% reduction in composite score (median difference –3.215; p = 1.71 × 10⁻⁵) (**Figure 2D**).

Unsupervised hierarchical clustering of the xCell enrichment scores of the CD45⁺ cell-type signatures separated samples largely by CD45⁺ depletion status. CD45⁺ depleted samples grouped broadly into separate branches characterized by lower z-scores across different cell type signatures. In contrast, non-depleted samples grouped together with broadly higher z-scores for the same cell-type features (**Figure 2E**).

### Differential gene expression analysis showed extensive transcriptional differences between high-grade and low-grade brain tumor patients and controls

To explore platelet gene expression differences between the groups, four pairwise comparisons were examined: 1: all brain tumor patients vs. controls (BTG vs. CTG), 2: high-grade tumor patients vs. controls (HGT vs. CTG), 3: high-grade tumor patients vs. low-grade tumor patients (HGT vs. LGT), and 4: low-grade tumor patients vs. controls (LGT vs. CTG). In the BTG vs. CTG comparison, a total of 398 genes were significantly differentially expressed, including 303 upregulated and 95 downregulated genes (**Figure 3A, Supplementary Table 1, Supplementary Figure 1)**. The most upregulated genes were *SLC6A19*, *CFAP77*, and *WFDC1*, whereas *RPL3L*, *SLC44A4*, and *ENSG00000227678* were the most downregulated.

**Figure 3.**
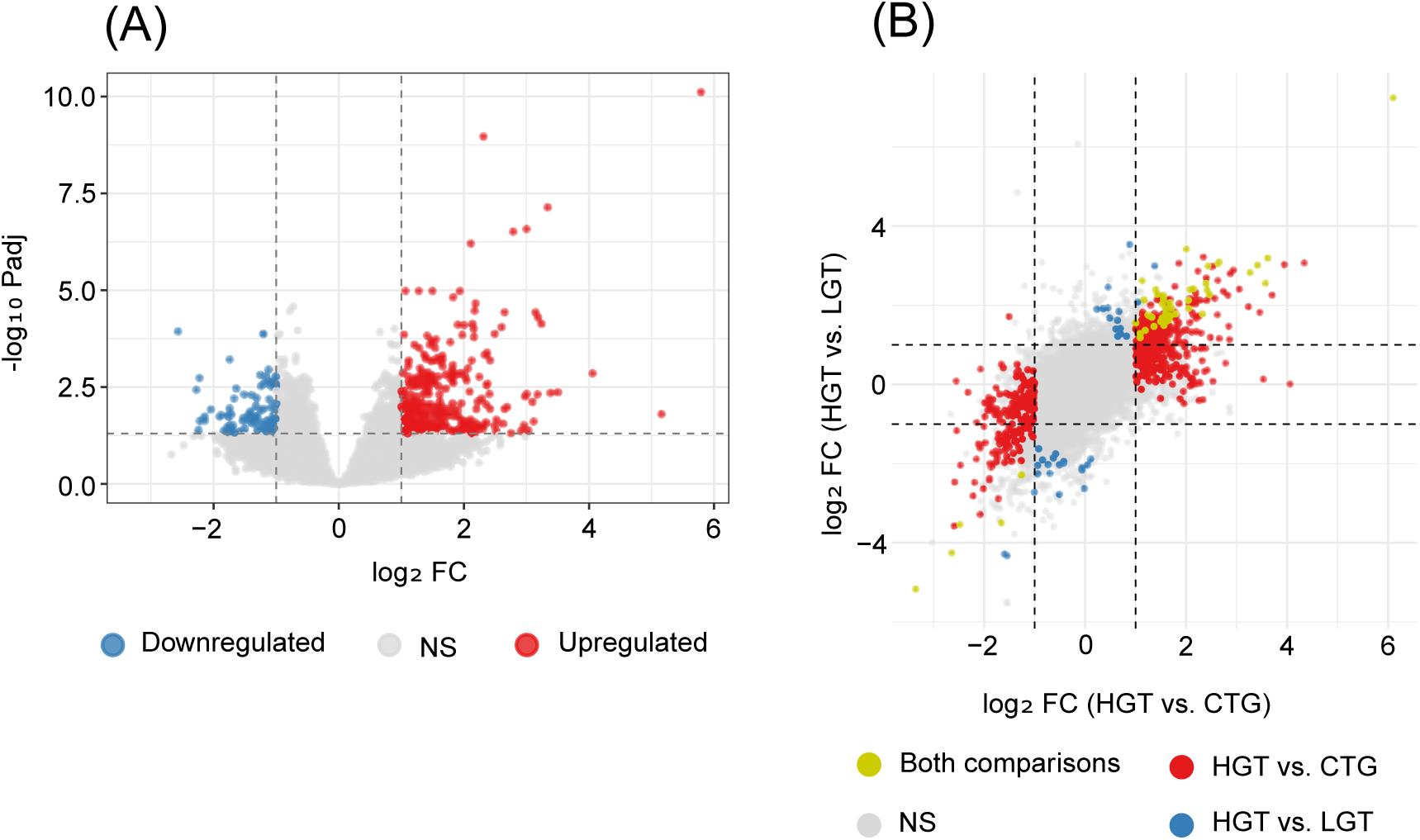
TEPs harbor altered transcriptomic profiles between different comparisons. **(A)** Volcano plot demonstrating DE analysis results between BTG vs. CTG illustrating up- and downregulated genes. **(B)** Four-way plot illustrating DE analysis results between HGT vs. LGT and HGT vs. CTG. Significance threshold FDR < 0.05 and an absolute log₂ fold change > 1. BTG = brain tumor group; CTG = cancer-free control group; HGT = high-grade tumor patients; LGT = low-grade tumor patients.

In the HGT vs. CTG comparison, 649 genes were significantly altered, of which 437 were upregulated and 212 downregulated (**Supplementary Table 2**). The top upregulated genes included *SLC6A19*, *CFAP77*, and *RPL29P26*, while *RN7SL769P*, *RN7SL8P*, and *NUP210L* showed the greatest downregulation.

When comparing samples from HGT vs. LGT, 85 genes were found to be differentially expressed (59 upregulated, 26 downregulated; **Supplementary Table 3**). The most upregulated genes were *SLC6A19*, *GPX1P1*, and *CCDC167*, whereas *OLFM4*, *ABCA13*, and *RN7SL674P* were the most downregulated. No genes met the significance threshold in the LGT vs. CTG comparison.

A summary of the relationships between the HGT vs. CTG and HGT vs. LGT results is presented in the four-way plot (**Figure 3B, Supplementary Figure 1**). Overall, HGT samples exhibited a greater number of transcriptional alterations compared to LGT and the CTG, indicating progressive molecular deregulation associated with systemic tumor involvement.

Individual assessment of the top 20 DE genes by adjusted p-value highlighted significant platelet gene expression differences between HGT, LGT, and CTG. In the HGT vs. CTG comparison, genes such as *S100A10*, *BIRC5*, *TUBG1, PLK1, SPAG5,* and *KIF20A* were markedly upregulated in HGT. None of the presented genes were markedly downregulated in the HGT **(Figure 4A).** In the HGT vs. LGT comparison, again, all presented genes were upregulated in the HGT group, e.g. *THEM5, NCEH1, MIR5703, GPX1P1* and *MRPL55.* No downregulation occurred here either **(Figure 4B).** Few genes overlapped in both comparisons, including *PSMB6, CCNB2, SLC6A10, RHCE, PTTG1* and *CDC20*.

**Figure 4.**
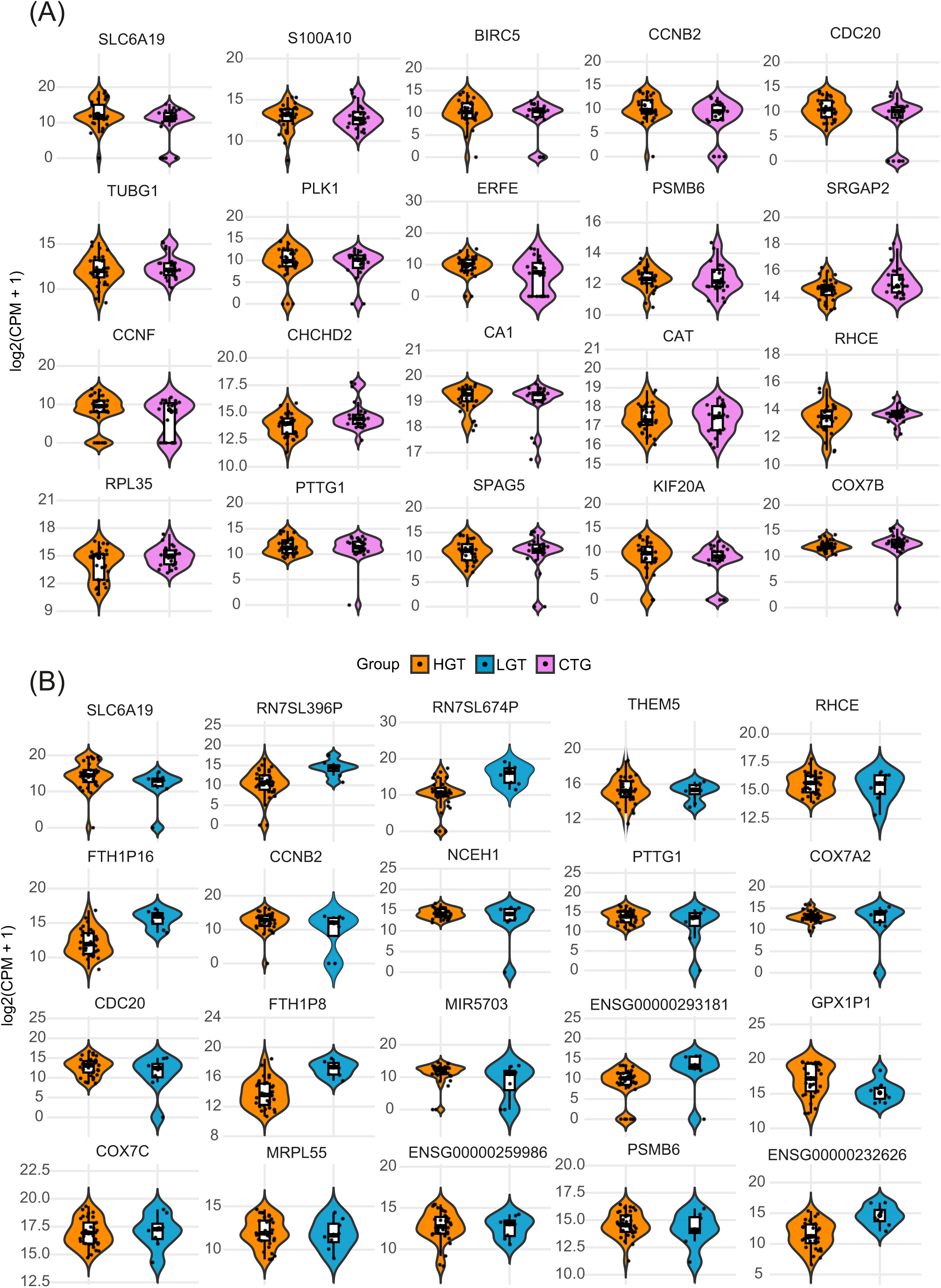
Top 20 most significantly differentially expressed genes between high-grade tumor patients, low-grade tumor patients, and cancer-free controls. **(A)** Violin and box plots illustrate normalized log₂(CPM + 1) expression values for each gene, highlighting distribution differences between HGT and CTG. Genes are ordered by adjusted *p*-value from DESeq2. HGT = high-grade tumor patients; CTG = cancer-free control group. **(B)** Violin and box plots show normalized log₂(CPM + 1) expression values for each gene, illustrating distribution differences between HGT and LGT. Genes are ranked by adjusted *p*-value from DESeq2. HGT = high-grade tumor patients; LGT = low-grade tumor patients.

To assess whether the overlapping genes in the group comparisons shifted in any direction, an effect-size concordance analysis was performed using the overlapping genes from the DE group comparison. All the overlapping genes (*PSMB6, PTTG1, RHCE, CCNB2, CDC20,* and *SLC6A19)* clustered near the diagonal line and exhibited concordant effects, indicating that they were consistently upregulated in HGT samples relative to both CTG and LGT. **(Supplementary Figure 2).**

### Discriminative power assessment of robust key platelet transcripts differentiating tumor groups and controls

To evaluate the ability of individual genes to distinguish between brain tumor patients and controls, ROC/AUC analysis was conducted on the same top 20 differentially expressed genes by adjusted p-value. The assessment revealed several platelet-expressed genes with strong discriminatory power between HGT and CTG. The leading candidates included *BIRC5* (AUC = 0.91), *TUBG1* (AUC = 0.90), *S100A10* (AUC = 0.86), *CDC20* (AUC = 0.85), and *KIF20A* (AUC = 0.85) **(Figure 5A).** When comparing HGT to LGT, the strongest classifiers were *MIR5703* (AUC = 0.95), *MRPL55* (AUC = 0.93), *COX7C* (AUC = 0.86), *NCEH1* (AUC = 0.84), and *COX7A2* (AUC = 0.81) **(Figure 5B).**

**Figure 5.**
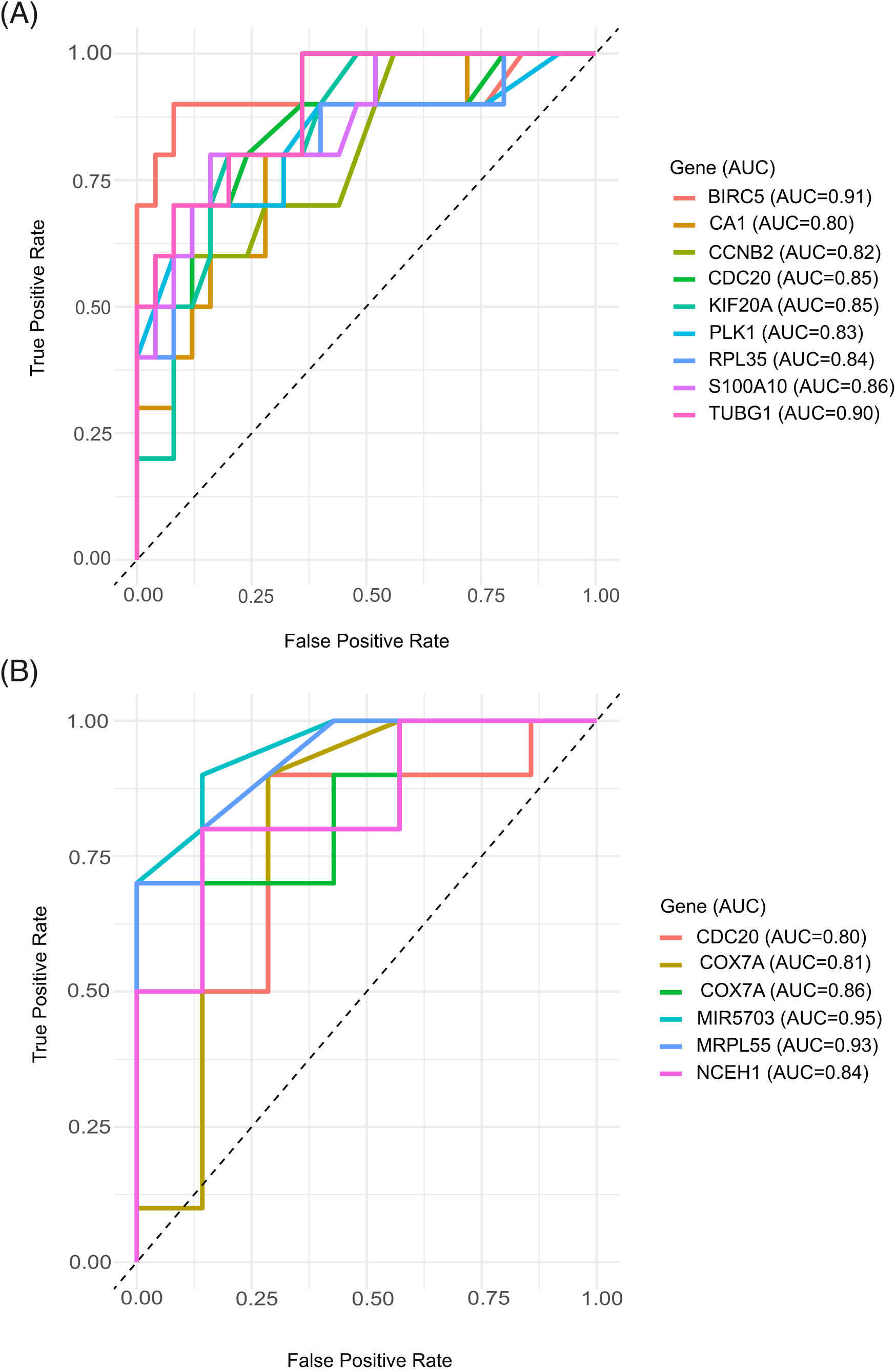
ROC analysis of transcriptomic biomarkers distinguishing high-grade tumor patients from low-grade tumor patients and controls. **(A)** ROC curve analyses showing the classification performance of transcriptomic biomarkers differentiating high-grade tumor patients from controls with an AUC value of at least 0.80. The ROC curves illustrate each gene’s discriminative capacity, with higher AUC values indicating stronger separation between HGT and CTG samples. The specific AUC value and the adjusted *p-*value are labeled within brackets for each gene. AUC=Area Under the Curve. HGT = high-grade tumor patients; CTG = cancer-free control group. **(B)** ROC analyses for genes differentially expressed between high-grade and low-grade tumor patients with AUC values at least 0.80. Each ROC curve demonstrates the sensitivity–specificity tradeoff for distinguishing tumor grades based on individual gene expression levels. The specific AUC value and the adjusted p-value are labeled within brackets for each gene. AUC=Area Under the Curve.

Heatmaps of the top 20 genes showed clear grouping. In HGT vs. CTG, several genes (e.g., *BIRC5, PLK1, CDC20, KIF20A, CCNB2, PTTG1*) were strongly upregulated in HGT platelets, while some controls showed downregulation of *CA1, CHCHD2*, and *CAT*. In HGT vs. LGT, distinct separation was again observed, with *MIR5703, MRPL55, SLC6A19, CCNB2,* and *COX7C* elevated in HGT, and *FTH1P8, FTH1P16, FKBP8, RN7SL674P,* and *RN7SL396P* higher in LGT/CTG (Supplementary Figures 2–3).

### Functional enrichment reveals mitotic and mitochondrial pathway dysregulation in high-grade tumor patients

To functionally interpret the genes represented in the top 20 genes from the DE analysis from both group comparisons, an over-representation analysis was performed. GO Biological Process enrichment revealed several statistically significant categories, predominantly related to cell division and mitotic regulation, chromosomal segregation, and mitochondrial processes.

Among the top enriched biological processes were nuclear division, chromosome segregation, organelle fission, mitotic spindle organization, and sister chromatid segregation, all of which are central to mitotic and meiotic cell division. In addition, several mitochondrial processes were significantly enriched, including mitochondrial electron transport, respiratory electron transport chain, ATP synthesis coupled electron transport, cellular respiration, and aerobic respiration **(Figure 6A).**

**Figure 6.**
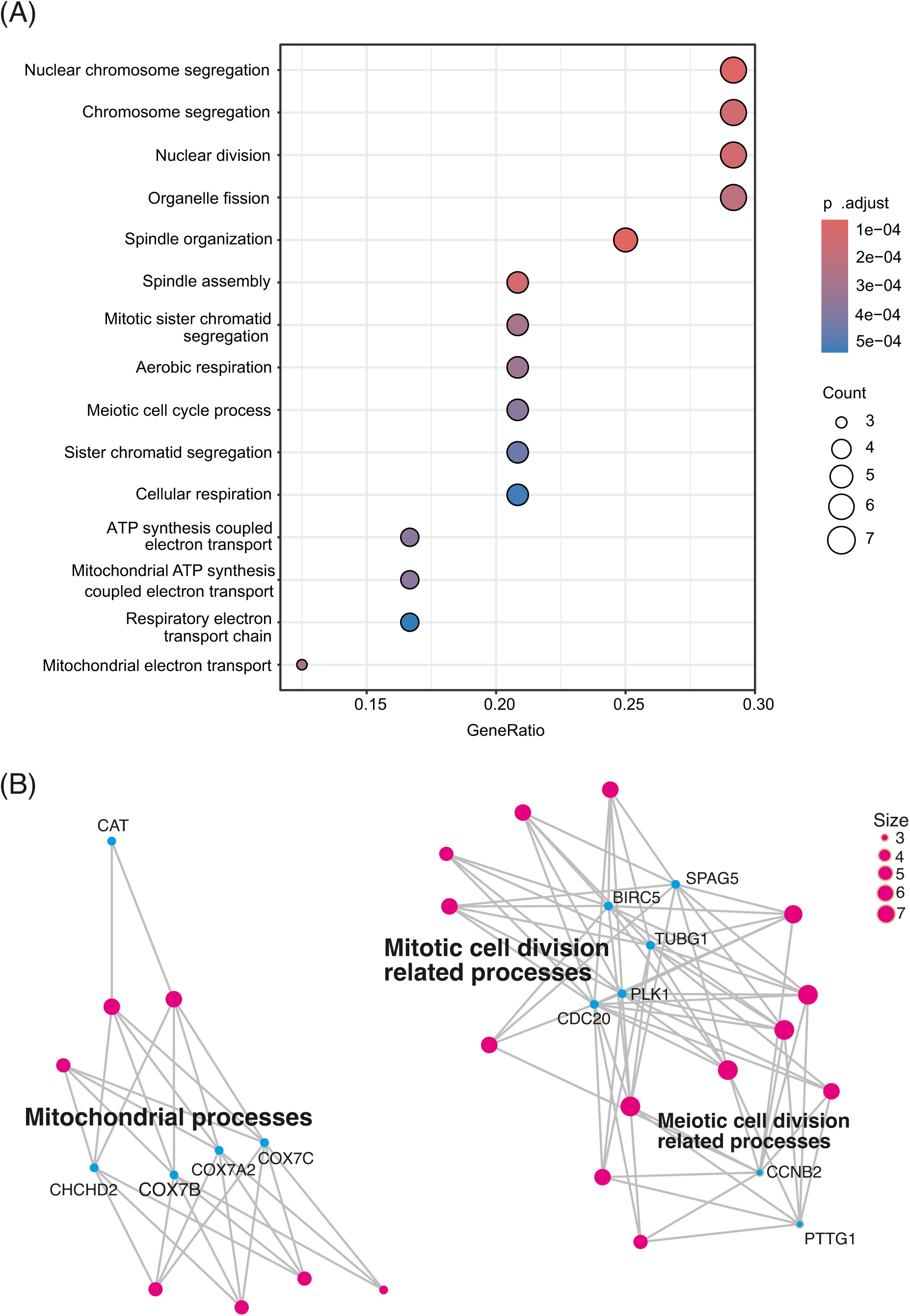
Over-representation analysis of the top 20 differentially expressed TEP transcriptomic signatures from DE analysis. **(A)** Functional enrichment analysis of the top 20 DE genes from HGT vs. LGT and HGT vs. CTG comparisons revealed that the largest and most enriched pathways were associated with nuclear division, organelle fission and chromosome segregation. The x-axis represents the gene ratio, and color indicates the adjusted p-value. **(B)** Pathways related to mitotic cell division were among the most significantly enriched, together with mitochondrial system regulation, reflecting both proliferative and metabolic gene alterations in TEPs.

The Gene-Concept Network plot **(Figure 6B)** visualized the relationships between enriched biological processes and their associated genes, highlighting key multifunctional nodes such as *CDC20, CCNB2, PTTG1, PLK1, BIRC5, SPAG5,* and *TUBG1.* These genes were shared among multiple enriched processes, indicating a central role in mitotic and meiotic control. The second cluster represents mitochondrial processes, such as mitochondrial electron transport, respiratory electron transport chain, and ATP synthesis coupled electron transport, primarily associated with *COX7C, COX7A2, COX7B,* and *CHCHD2.* The strong network connectivity underscores the convergence of proliferative and metabolic pathways characteristic of high-grade tumors.

Studying all the differentially expressed genes from BTG vs. CTG comparison, similar pathways were pronounced, including several pathways of the mitotic cell division **(Supplementary Figure 4).** All recognized pathways in this comparison are listed in the **Supplementary Table 1.**

### Binary logistic regression model validated the predictive power of the platelet gene expression signature for high-grade tumor patients

A binomial elastic-net logistic regression model with patient-grouped cross-validation was used to assess whether TEP-derived gene expression could distinguish high-grade tumor patients from cancer-free controls. The model performed robustly, achieving a cross-validated AUC of 0.89 **(Figure 7A)** and clear separation of predicted and true sample labels **(Figure 7B)**. Out-of-fold performance metrics **(Table 3)** showed a sensitivity of 79%, a specificity of 80%, and an overall accuracy of 79%, confirming that the selected genes form a reproducible high-grade tumor signature. Coefficient analysis identified key contributors to the classification, including *S100A10, CA1, TUBG1, CDC20, CCNB2, KIF20A, COX7C,* and *COX7A2*, while *RPL35* and *PLK1* provided weaker discrimination **(Figure 7C)**.

**Figure 7.**
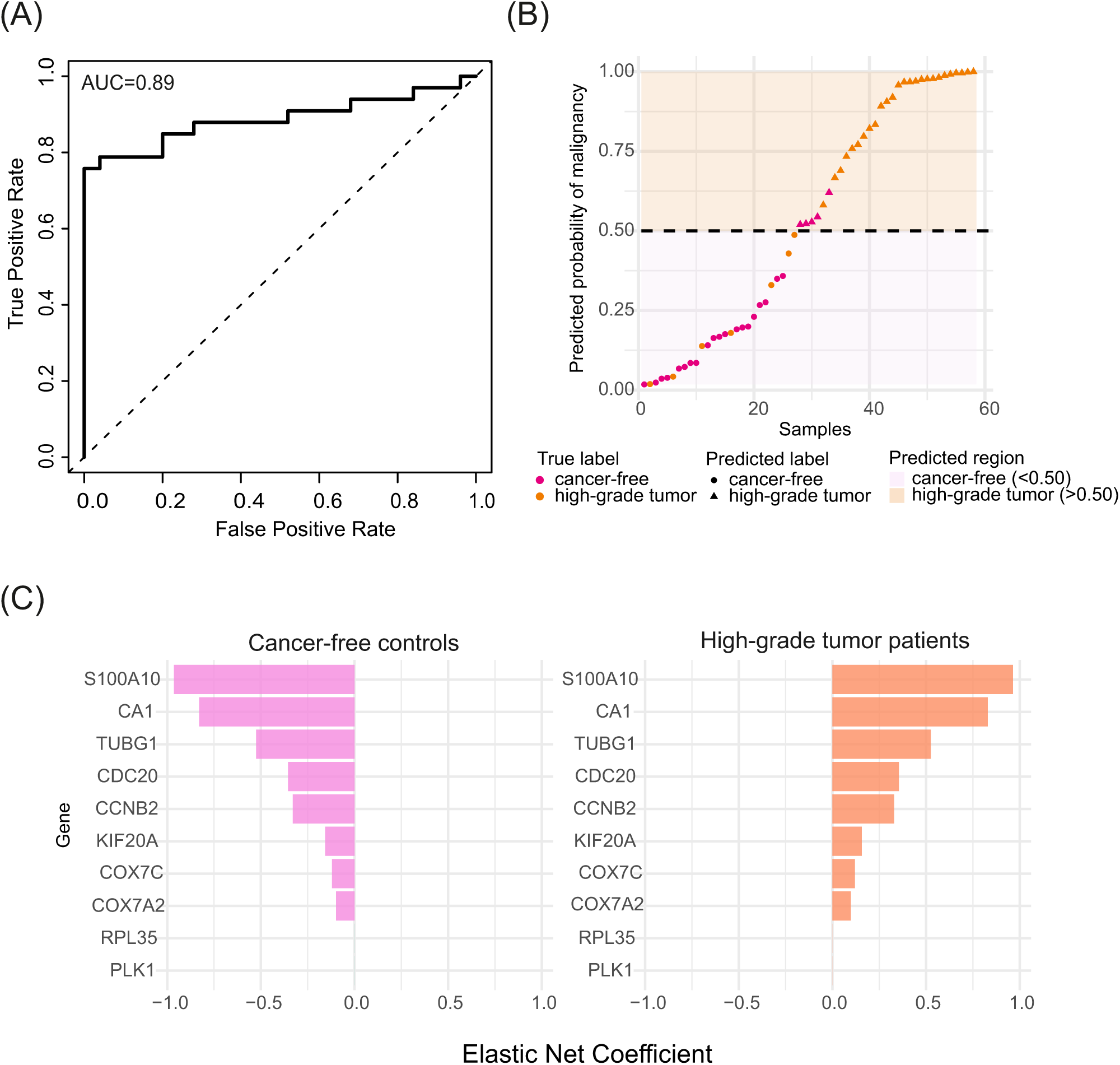
Logistic regression model distinguishes controls and high-grade tumor patients. **(A)** ROC curves illustrating the performance of the logistic regression model in distinguishing between HGT and CTG samples. The AUC value reached 0.89, indicating a promising overall discriminative capability of the model. **(B)** Each point represents an individual sample. The y-axis shows the predicted probability of having a high-grade tumor patient from the logistic regression model, and the x-axis shows the sample index after sorting by predicted probability (low to high). The background shading indicates the model’s predicted class region, with violet corresponding to predicted *cancer-free control* (probability < 0.5) and orange corresponding to predicted *high-grade tumor patient* (probability ≥ 0.5). The dashed horizontal line at 0.5 marks the classification threshold. Point color denotes the true class (cancer-free control or high-grade tumor patient), and point shape denotes the predicted class, allowing misclassified samples to be visually identified where color and shape disagree. **(C)** Coefficient weights from the final elastic-net binomial logistic regression model are shown for genes with non-zero contributions to classification. Positive coefficients correspond to genes enriched in high-grade tumor patients, while negative coefficients represent their downregulation in cancer-free controls.

**Table 3.** Performance metrics of the logistic regression model. Summary of results for the logistic regression classifier, including counts of true/false positives and negatives and performance metrics per class.

| Class | TP | FP | TN | FN | Sensitivity | Specificity | AUC | Accuracy |
| --- | --- | --- | --- | --- | --- | --- | --- | --- |
| CTG | 20 | 7 | 26 | 5 | 0.80 | 0.79 | 0.89 | NA |
| HGT | 26 | 5 | 20 | 7 | 0.79 | 0.80 | 0.89 | NA |
| Overall | 46 | 12 | NA | NA | NA | NA | NA | 0.79 |

Abbreviations: TP = true positive; FP = false positive; TN = true negative; FN = false negative; AUC = area under the ROC curve. Sensitivity = $TP/(TP+FN)$ ; Specificity = $TN/(TN+FP)$ .

## Discussion

This study investigated the transcriptomic landscape of TEPs in pediatric brain tumors using sequencing of platelet RNA from HGT, LGT, and CTG. Our findings demonstrated the feasibility and analytical robustness of platelet-derived RNA profiling in pediatric brain tumors based on plasma collected at diagnosis and during therapy and follow-up. By integrating multiple group comparisons and biomarker identification approaches, we showed that platelet RNA profiles can effectively distinguish both the presence and nature of brain tumors. Collectively, these results provide novel insights into platelet RNA biology in patients with childhood brain tumors and support the potential of TEP signatures as a minimally invasive biomarker source for malignancy detection and monitoring. Although similar results have been demonstrated in the adult population, as far as we know, this is the first reported study for the use of TEPs in pediatric cancer patients.

Many brain tumor biomarkers fail to enter the bloodstream because of the blood–brain barrier, limiting the clinical utility of several liquid biopsy approaches, including circulating tumor DNA, circulating tumor cells, and extracellular vesicles. In contrast, TEPs can actively interact with brain tumors and transport tumor-derived molecular information across the blood-brain barrier (11). This ability allows TEPs to capture dynamic tumor signals that are often undetectable with other blood-based liquid biopsy methods, making them an attractive tool for the detection and monitoring of brain tumors (18).

Both platelet count and RNA load vary in response to physiological and pathological processes. To assess the suitability of our samples for downstream transcriptomic analyses, we first evaluated RNA yield and integrity across all samples. Despite the inherent variability in platelet RNA content, the overall RNA quality was high. This indicates that pre-analytical procedures, including rapid sample processing and standardized isolation, preserved RNA integrity. Importantly, platelet count did not correlate with RNA yield, consistent with previous reports in adult cohorts (11). This finding reinforces that the amount of extractable platelet RNA is influenced more by other factors than platelet count alone, such as the immature platelet fraction and clinical states (e.g., sepsis) (28,29). Consequently, reliable RNA extraction and downstream analyses are feasible even in samples from thrombocytopenic patients, which is relevant for pediatric oncology, where platelet counts may fluctuate due to therapy or disease. However, in cytopenic patients, it is important to consider the effect of platelet infusions that they might receive during therapy. Infusions of donor-derived platelets could potentially lead to false-negative results in TEP transcriptomic analyses, as the infused platelets may not carry the patient’s tumor-associated RNA signatures. Given the short lifespan of platelets, the transfused cells would be rapidly replaced by newly produced platelets originating from the patient’s own bone marrow. It is also conceivable that transfused donor platelets might undergo tumor education after infusion, acquiring tumor-associated transcripts despite their external origin. Nevertheless, there remains a significant gap in knowledge regarding how rapidly platelets can be reprogrammed *in vivo*, underscoring the need for dedicated experimental studies to address this in the future.

Many previous platelet transcriptomic studies have relied solely on double-centrifugation protocols, which may not fully eliminate residual leukocytes (30–32). Leukocyte contamination is a possible confounding factor in platelet RNA studies, as even minimal numbers of leukocytes contribute disproportionately to total RNA and can distort transcriptomic readouts (33). Our comparison between CD45⁺ depleted and non-depleted samples confirmed that depletion effectively reduced leukocyte-derived transcripts, as evidenced by significantly lower composite xCell scores in depleted samples. The clustering of non-depleted samples further highlights the substantial transcriptomic shift introduced by leukocyte contamination. These findings suggest the CD45⁺ depletion approach as an essential step for improving platelet RNA purity and subsequent downstream analyses, particularly in plasma samples with variable leukocyte content.

Platelets from brain tumor patients exhibited substantial transcriptomic dysregulation when compared with cancer-free controls, strongly suggesting that the presence of a tumor induces systemic platelet reprogramming. Importantly, the number of altered transcripts was the highest in high-grade tumor patient vs. cancer-free control patient comparison, suggesting that the extent of platelet remodeling scales with the presence of malignancy. Together, these patterns support the concept that platelet transcriptomic changes reflect both tumor presence and malignancy, consistent with the framework of tumor-educated platelets. This aligns with previous studies, where non-metastatic tumor patients have been compared to metastatic tumor patients (18).

Interestingly, no significant transcriptional differences were observed between low-grade tumor patient samples and control patient samples. This may indicate that low-grade brain tumors exert a weaker systemic influence on platelet RNA remodeling. Alternatively, the absence of detectable differences could reflect the modest sample size, heterogeneity of tumor subtypes, or limited sensitivity of bulk platelet RNA-seq to subtle molecular alterations in LGT. This finding supports the notion that patients with low-grade pediatric tumors share greater transcriptomic similarity of platelets with cancer-free controls, consistent with their slower progression rate and lower systemic activity (34).

Assessment of the most differentially expressed genes in HGT samples revealed multiple transcripts with known roles in cancer pathogenesis. Several genes (*SLC6A19, PSMB6, CDC20, CCNB2, SLC6A19, RHCE, PTTG1*) were consistently dysregulated in both group comparisons (HGT vs. CTG and HGT vs. LGT). Out of these, *PSMB6, CCNB2, CDC20* and *PTTG1* have been associated with diverse oncogenic processes such as cell cycle deregulation and chromosomal disorganization across tumor types (35–37), while *SLC6A19* has been linked to cancer pathogenesis in renal malignancies (38). The strong concordance of these overlapping genes supports their robustness as TEP markers.

In the platelet gene expression comparison of HGT vs. CTG, we identified several additional cancer-associated transcripts, such as *TUBG1, SPAG5, BIRC5, S100A10, KIF20A, RPL35, PLK1* and *CA1.* These genes have been broadly associated with tumor proliferation, cell-cycle dysregulation, and poor clinical outcomes in patient cohorts with elevated expression (39–46). Similarly, the HGT vs. LGT comparison revealed differential expression of tumor-related genes such as *MRPL55, COX7C, THEM5, NCEH1, MIR5703,* and *COX7A*, all previously implicated in cancer progression (41,47–52). To be noted, nine genes identified in the HGT vs. CTG comparison and six genes from the HGT vs. LGT comparison demonstrated significant discriminative power in our classification assessment. This convergence between differential expression and predictive performance further supports the diagnostic potential of these genes as TEP-derived biomarkers for pediatric brain tumors.

Building on these findings, functional enrichment pointed to strong associations with mitotic cell division, spindle organization, and chromosome segregation. These processes are normally restricted to proliferating nucleated cells and are therefore unexpected in anucleate platelets. Mitochondrial pathways were also enriched, potentially reflecting oxidative stress and tumor-driven metabolic changes (53). Another theory for this is that platelets may uptake tumor-derived mitochondria, which can later be transferred to new tumor cells to enhance their energy production and survival (54). The presence of these signatures in platelets is striking, given their lack of a nucleus and usual physiological role, suggesting that these alterations likely represent true cancer-related biology rather than incidental signals. Future dedicated experimental models will be needed to clarify how platelets acquire and potentially support these processes in pediatric brain tumors.

To enable the utilization of platelet RNA sequencing in daily clinical practice, larger datasets and the development of a validation model for sequencing data would be necessary in the future. We created a preliminary model aiming to distinguish CTG from HGT. The binary logistic regression model achieved promising results and recognized the CTG from HGT with 80% sensitivity. These results emphasize that identifying relevant biomarkers for each group comparison and optimizing model training are both essential for reliable model development.

In the individual sample scoring by the validation model, some noteworthy patterns emerged. For some patients, both diagnostic and follow-up samples during therapy were available. Some of the follow-up samples were collected when remission was achieved during therapy. Therefore, these samples should likely group closer to the CTG samples instead of the HGT samples, and that was also noticed in the model classification for a few samples. Such observation affects the accuracy of the model, emphasizing the importance of considering patient metadata when interpreting model outcomes, as clinical context critically influences the reliability of classification results.

While this study provides valuable insights into the platelet RNA profiles of pediatric brain tumor patients, a few limitations should be acknowledged. First, the small sample size may limit the generalizability of the findings, a common limitation in pediatric oncology studies when studying rare diseases. Future studies with larger cohorts are warranted to validate the results and explore the potential clinical implications of the identified gene expression patterns. The study was also conducted as a single-center study, possibly limiting the generalizability of the patient population and risking institutional bias. In this study, we also studied a large variability of tumors, which may also affect the results. Studies including more narrowly specific tumor diagnoses could improve the precision of analyses. Additionally, broader longitudinal studies may be necessary to further investigate the dynamics of platelet RNA profiles in the context of disease progression and treatment response. Assessing the validation model, the logistic regression analysis was not trained enough for observation of clinical status and treatment results, leading to mislabeling, which affects both the sensitivity and specificity. Although these observations should be kept in mind, this study already provides a broad insight into platelet transcriptomics in pediatric brain tumor patients.

## Conclusion

In conclusion, our findings demonstrated the feasibility and biological relevance of platelet RNA sequencing as a minimally invasive liquid biopsy approach for pediatric brain tumors. Platelet transcriptomes robustly distinguished patients with high-grade tumors from patients with low-grade tumors and cancer-free controls, indicating that platelet RNA profiles reflect both tumor presence and clinical severity. The identified differentially expressed genes and enriched pathways support substantial tumor-driven reprogramming consistent with the concept of tumor-educated platelets. The preliminary classification model further demonstrated the diagnostic potential of TEP-derived signatures, although further optimization and larger datasets are required to enhance its performance. While sample size and biological heterogeneity pose limitations, these findings provide a foundation for the development of TEP-based diagnostics for pediatric brain tumors.

### Abbreviations

AUC: Area under the curve
BTG: brain tumor patient group
CNS: central nervous system
CTG: control group; pediatric patients with no known cancer history
DE: Differential gene expression
FDR: false discovery rate
GO: Gene Ontology
HGT: high-grade tumor patients
LGT: low-grade tumor patients
MRI: magnetic resonance imaging
NS: not significant
PCA: Principal component analysis
RIN: RNA integrity
ROC: receiver operating characteristic
RPKM: reads per kilobase per million
RT: room temperature
TEP: tumor-educated platelet

## Author contributions

KE & VP conceived the study. MT, AH, PV, VP & KE participated in the design of study. MT was responsible for sample processing and RNA-sequencing. MT, AH, JH, MK & EV performed data analyses and investigations. AH, SL, MWO, AMS, RN, PI, PV, AK, PKP, APK, NV, JS, OS, JK, OT & VP contributed to patient sample and/or clinical data acquisition. SK, AKA, VP & KE provided resources and supervised the study. MT & AH wrote the first manuscript draft. All authors revised the manuscript and approved the final submitted version.

## Acknowledgements

The prospective sample collection was carried out in collaboration with Helsinki Biobank. We thank all study participants for their generous participation in Helsinki Biobank and this study. Biostatistics Consulting Service at the University of Helsinki and Helsinki University Hospital is acknowledged for their support in data analyses. We express our gratitude to all nurses, physicians, and other hospital staff at New Children’s Hospital (Helsinki, Finland) whose flexibility and help have enabled smooth patient recruitment and sample collection.

## Ethical statement

This study was conducted in line with the principles of the Declaration of Helsinki. The study was approved by the Regional Ethics Committee of the Helsinki University Hospital (HUS/2124/2023). Written informed consent was obtained from all patients and/or their parents/legal guardians. All samples were collected during diagnostic procedures to avoid redundant sampling.

## Financial statement

This study was supported by Väre Foundation for Pediatric Cancer Research (KE), AAMU Pediatric Cancer Foundation (VP), Finnish Pediatric Research Foundation (KE), Päivikki and Sakari Sohlberg Foundation (KE), Biomedicum Helsinki Foundation (AH), Helsinki University Hospital Research Grants (AKA), and iCANDOC Doctoral Education Pilot in Precision Cancer Medicine (AH).

## Data availability

The datasets used during the current study are available from the corresponding author on reasonable request.

---

**Supplementary Table 1.** Gene Ontology (GO) biological process over-representation analysis results for all differentially expressed genes between brain tumor patients and control patients. The table lists enriched GO terms, associated genes, gene counts, gene ratios, and adjusted *p*-values.

**Supplementary Figure 1**. Differentially expressed platelet genes across diagnostic group comparisons. Barplot showing the number of significantly upregulated and downregulated genes in comparisons between (i) HGT vs. CTG, (ii) HGT vs. LGT, and (iii) BTG vs. CTG. Bars represent the count of genes meeting the differential expression threshold (FDR < 0.05 and |log₂FC| > 1). CTG = cancer-free control group; BTG = Pediatric brain tumor patients; HGT = high-grade tumor patients; LGT = low-grade tumor patients.

**Supplementary Figure 2.** *Effect-size concordance of differentially expressed genes across high-grade tumor patient comparisons.* Scatterplot showing log₂ fold changes (log₂FC) for HGT versus CTG (y-axis) and HGT versus LGT (x-axis) comparisons. Each point represents a gene, with those near the diagonal exhibiting similar effect sizes and directions across both contrasts, indicating concordant differential expression. CTG = cancer-free control group; HGT = high-grade tumor patients; LGT = low-grade tumor patients.

**Supplementary Figure 3.***Heatmap of the top 20 genes comparing high-grade tumor patients and controls*. Heatmap showing expression patterns of top genes identified by filtering genes with the top 20 best adjusted p-values, distinguishing high-grade tumor patients from controls by DESeq2 analysis. Each column represents an individual sample, and each row a gene. Expression values are scaled per gene (row-wise Z-score). Metadata annotations above the heatmap indicate group (CTG, LGT, HGT), specific diagnosis, biological gender, and sampling timepoint (before or during therapy). Distinct clustering separates tumor and control groups, reflecting consistent differential expression among selected markers. CTG = cancer-free control group; HGT = high-grade tumor patients; LGT = low-grade tumor patients.

**Supplementary Figure 4.** *Heatmap of the top 20 genes comparing high-grade and low-grade tumor patients.* Expression heatmap of the top 20 genes with the best adjusted p-values from DESeq2 analysis between HGT and LGT samples. The accompanying metadata tracks group, diagnosis, biological gender, and timepoint of sampling, highlighting patient heterogeneity. Row-wise Z-scored expression levels demonstrate clear separation between tumor groups (HGT vs. LGT), indicating TEP expression signatures linked to the grade of the neoplasm. CTG = cancer-free control group; HGT = high-grade tumor patients; LGT = low-grade tumor patients.

**Supplementary Figure 5.** *ORA GO enrichment analysis of differentially expressed genes between the brain tumor group and controls.* (**A**) Gene Ontology (GO) over-representation analysis showing enriched biological processes among all studied genes. Dot size represents the number of genes per GO term, color indicates adjusted *p*-value, and the x-axis shows the gene ratio. Enriched terms are predominantly related to mitotic spindle assembly, spindle checkpoint signaling, and chromosome segregation, all belonging to the mitotic cell division process. (**B**) Category-gene network plot illustrating the relationships between enriched GO terms (pink nodes) and their associated genes (blue nodes), highlighting key genes involved in mitotic checkpoint regulation such as *BIRC5, CDC20, PLK1, CCNB1, MAD2L1,* and *SKA3*.

