## Supplementary Figures for "Platelet transcriptomic signatures in pediatric brain tumors distinguish cancer from cancer-free control"

Supplementary Figure 1

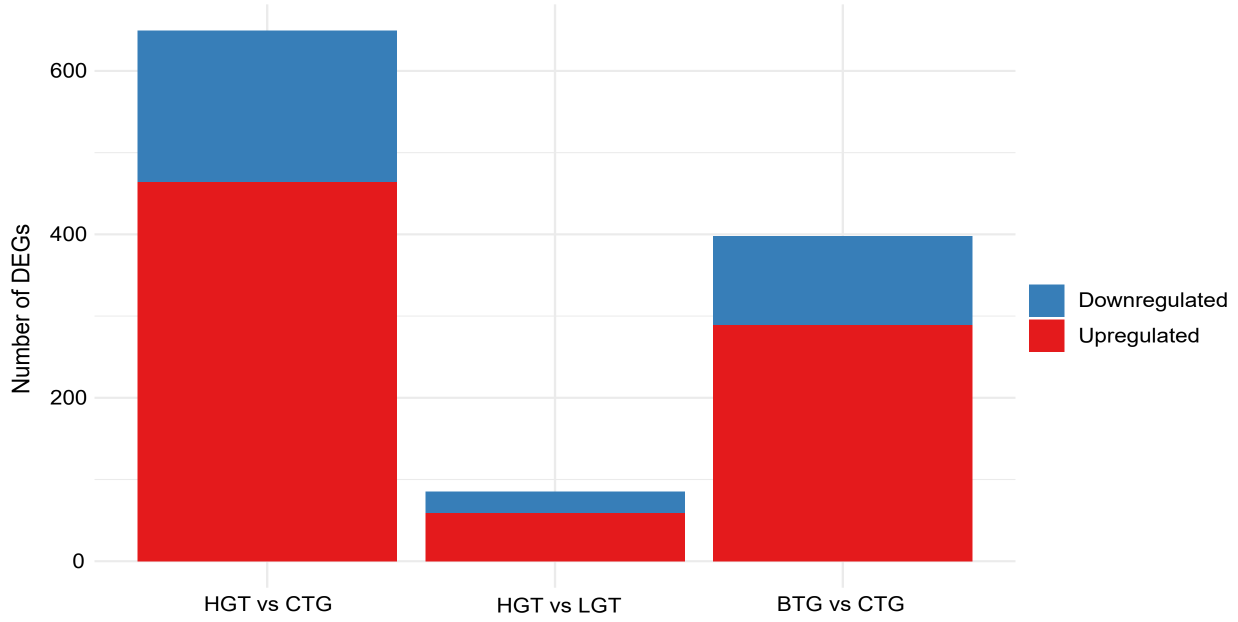


**Supplementary Figure 1.** *Differentially expressed platelet genes across diagnostic group comparisons.*

Barplot showing the number of significantly upregulated and downregulated genes in comparisons between (i) HGT vs. CTG, (ii) HGT vs. LGT, and (iii) BTG vs. CTG. Bars represent the count of genes meeting the differential expression threshold (FDR < 0.05 and $|\log_2 FC| > 1$). CTG = cancer-free control group; BTG = Pediatric brain tumor patients; HGT = high-grade tumor patients; LGT = low-grade tumor patients.

Supplementary Figure 2

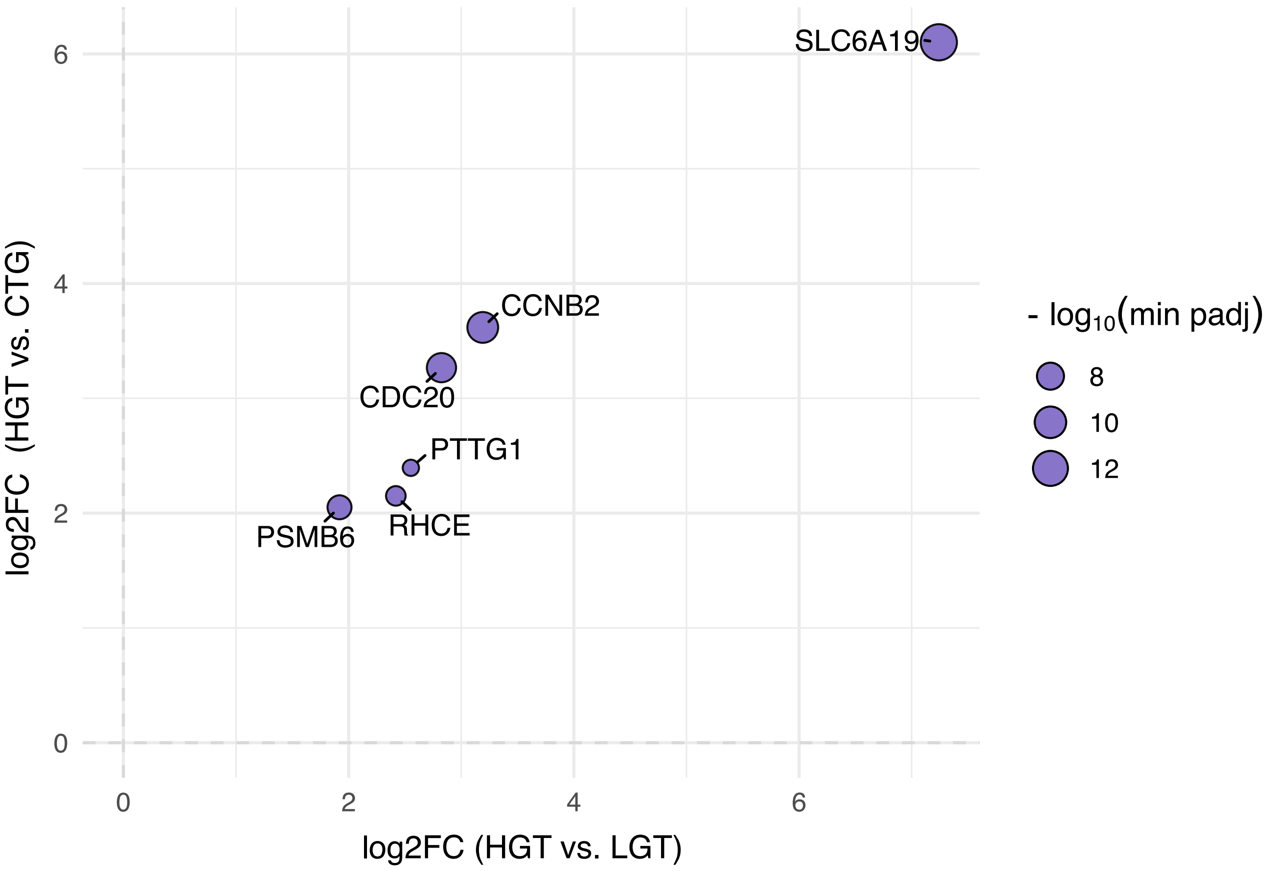


**Supplementary Figure 2.** *Effect-size concordance of differentially expressed genes across high-grade tumor patient comparisons.* Scatterplot showing $\log_2$ fold changes ($\log_2$FC) for HGT versus CTG (y-axis) and HGT versus LGT (x-axis) comparisons. Each point represents a gene, with those near the diagonal exhibiting similar effect sizes and directions across both contrasts, indicating concordant differential expression. CTG = cancer-free control group; HGT = high-grade tumor patients; LGT = low-grade tumor patients.

Supplementary Figure 3

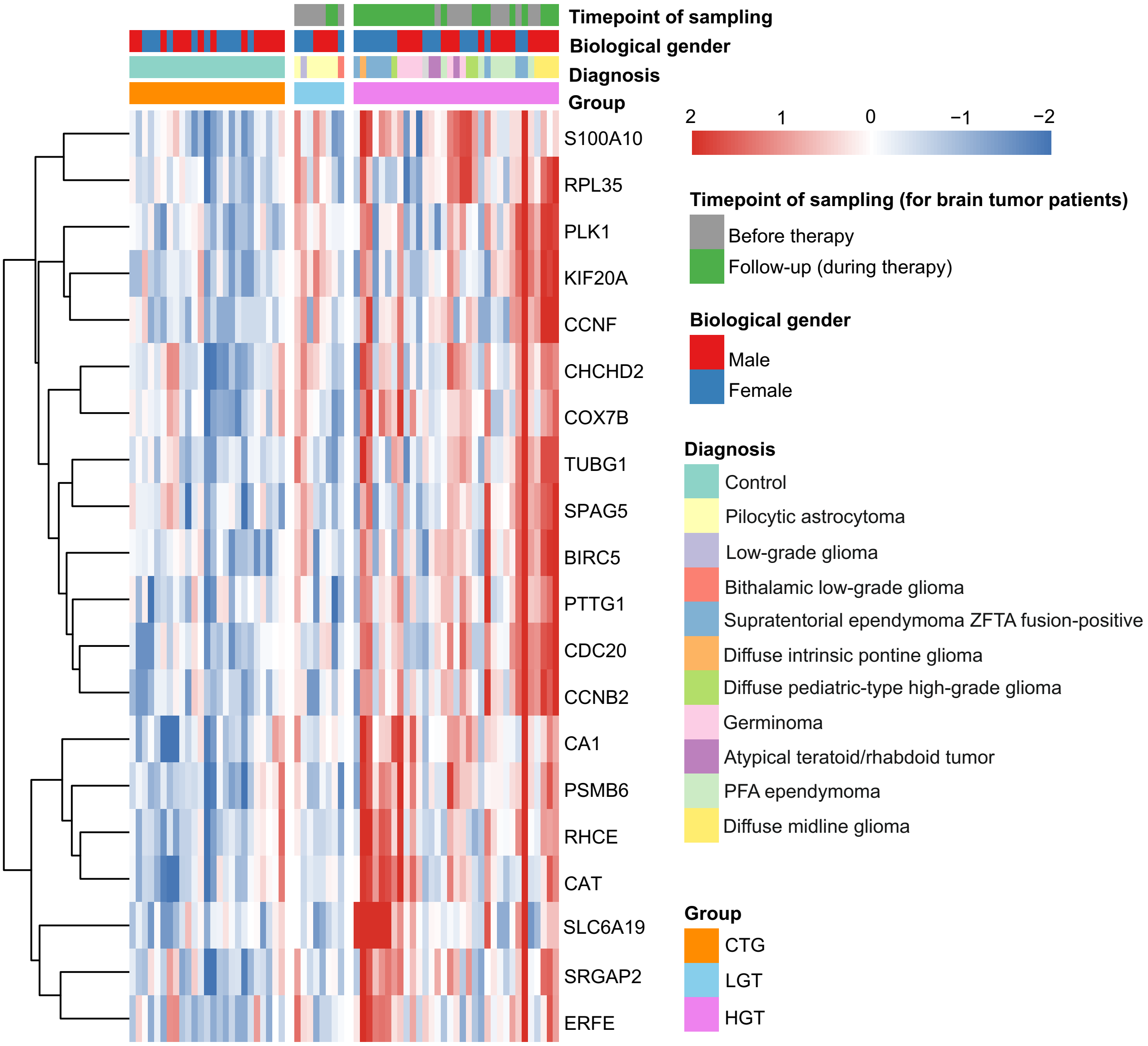


**Supplementary Figure 3.** *Heatmap of the top 20 genes comparing high-grade tumor patients and controls.*

Heatmap showing expression patterns of top genes identified by filtering genes with the top 20 best adjusted p-values, distinguishing high-grade tumor patients from controls by DESeq2 analysis. Each column represents an individual sample, and each row a gene. Expression values are scaled per gene (row-wise Z-score). Metadata annotations above the heatmap indicate group (CTG, LGT, HGT), specific diagnosis, biological gender, and sampling timepoint (before or during therapy). Distinct clustering separates tumor and control groups, reflecting consistent differential expression among selected markers. CTG = cancer-free control group; HGT = high-grade tumor patients; LGT = low-grade tumor patients.

Supplementary Figure 4

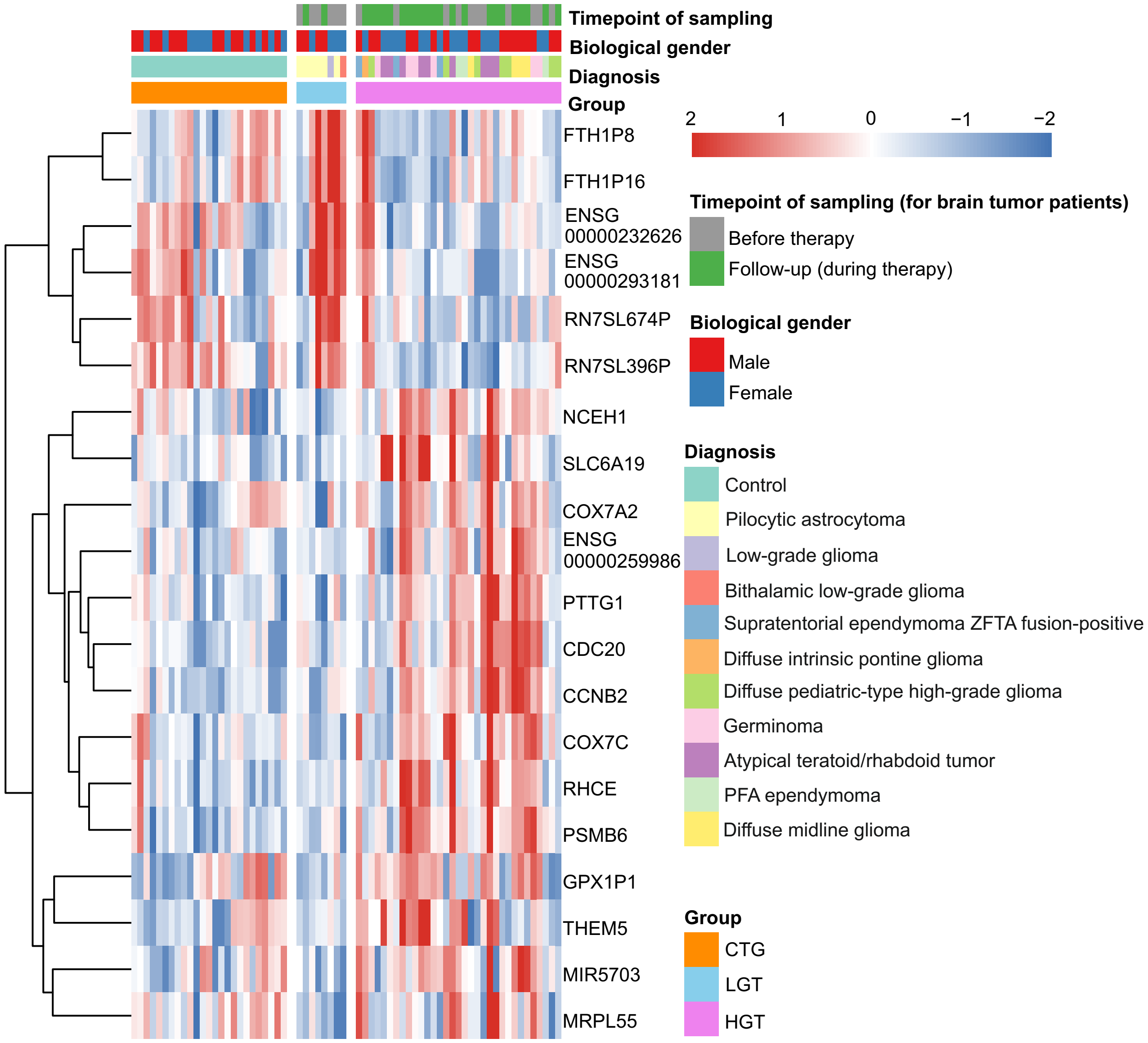


**Supplementary Figure 4.** *Heatmap of the top 20 genes comparing high-grade and low-grade tumor patients.*

Expression heatmap of the top 20 genes with the best adjusted p-values from DESeq2 analysis between HGT and LGT samples. The accompanying metadata tracks group, diagnosis, biological gender, and timepoint of sampling, highlighting patient heterogeneity. Row-wise Z-scored expression levels demonstrate clear separation between tumor groups (HGT vs. LGT), indicating TEP expression signatures linked to the grade of the neoplasm. CTG = cancer-free control group; HGT = high-grade tumor patients; LGT = low-grade tumor patients.

Supplementary Figure 5

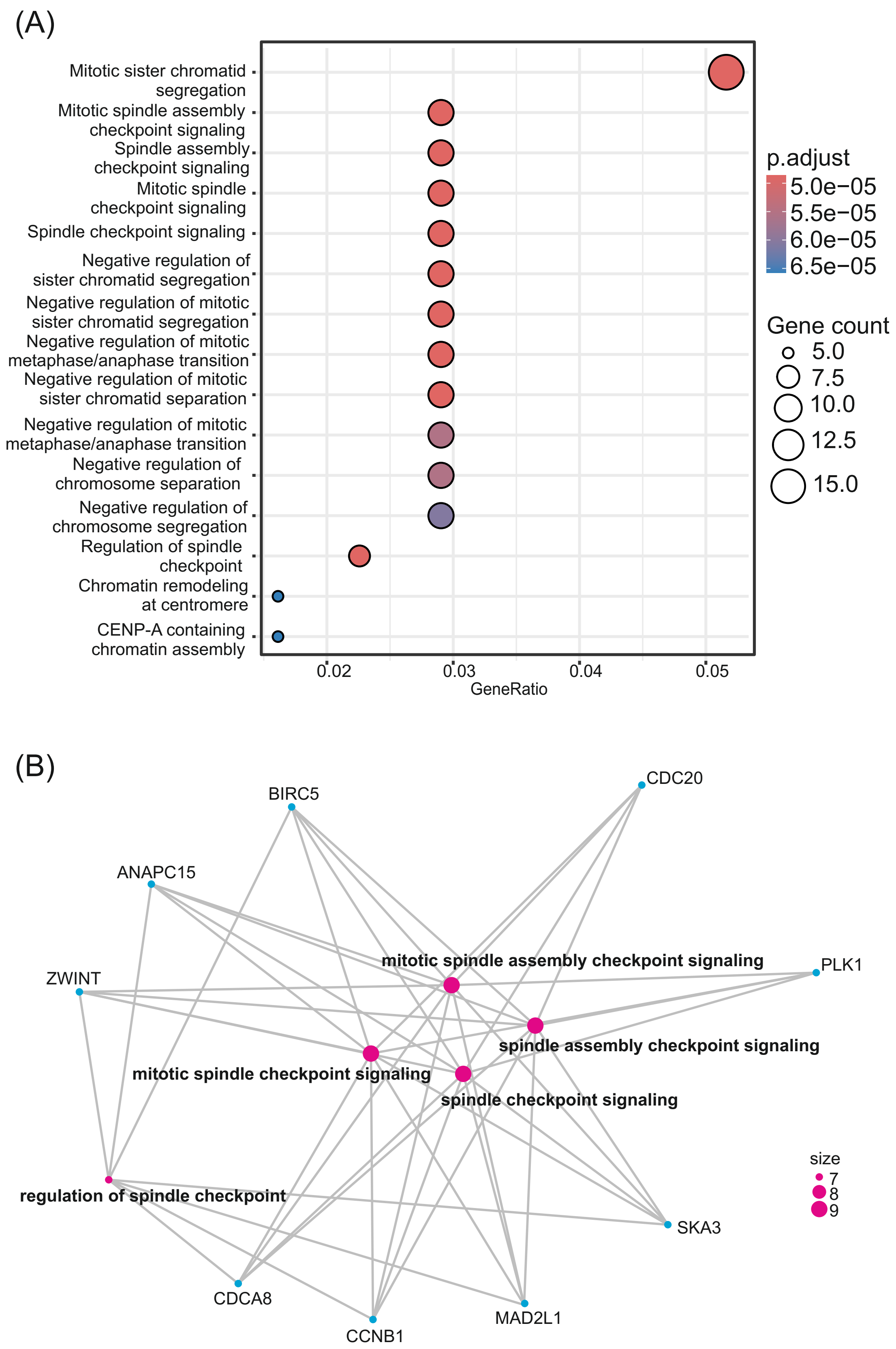


**Supplementary Figure 5. ORA GO enrichment analysis of differentially expressed genes between the brain tumor group and controls.** (A) Gene Ontology (GO) over-representation analysis showing enriched biological processes among all studied genes. Dot size represents the number of genes per GO term, color indicates adjusted *p*-value, and the x-axis shows the gene ratio. Enriched terms are predominantly related to mitotic spindle assembly, spindle checkpoint signaling, and chromosome segregation, all belonging to the mitotic cell division process. (B) Category-gene network plot illustrating the relationships between enriched GO terms (pink nodes) and their associated genes (blue nodes), highlighting key genes involved in mitotic checkpoint regulation such as *BIRC5*, *CDC20*, *PLK1*, *CCNB1*, *MAD2L1*, and *SKA3*.
